# A Selective Increase in OC Symptoms is Driving Information Seeking and Guideline Adherence During the Covid-19 Pandemic

**DOI:** 10.1101/2020.12.08.20245803

**Authors:** Alisa M. Loosen, Vasilisa Skvortsova, Tobias U. Hauser

## Abstract

**Background:** Increased mental health problems as a reaction to stressful life events, such as the Covid-19 pandemic, are common. Critically, successful adaptation helps reduce such symptoms to baseline, preventing long-term psychiatric disorders. It is thus important to understand whether and which psychiatric symptoms only show transient elevations, and which persist long-term and become chronically heightened. At particular risk for the latter trajectory are disorders with symptoms directly affected by the pandemic, such as obsessive-compulsive disorder.

**Methods:** In this longitudinal large-scale study (N=416), we assessed how obsessive-compulsive (OC), anxiety, and depression symptoms changed throughout the course of the first pandemic wave in a sample of the general UK public. We further examined how these symptoms affected pandemic-related information seeking and adherence to governmental guidelines.

**Findings:** All psychiatric domains were initially elevated, but showed distinct adaptation patterns. Depression scores decreased during the first pandemic wave, however, OC symptoms further increased, even after the end of lockdown. These OC symptoms were directly linked to Covid-related information seeking which gave rise to higher adherence to government guidelines.

**Interpretation:** The rise and persistence of OC symptoms, despite the ease of Covid-19 restrictions, shows that OCD is disproportionally and chronically affected by the pandemic. This is particularly worrying with regards to the long-term impact of the Covid-19 pandemic on public mental health and indicates that patients with OCD may require particular treatment efforts.

## Introduction

Humans are innately averse to uncertainty. When uncertain, stress levels increase and peoples’ mental health can suffer (1,2). The global coronavirus SARS-CoV-2 (Covid-19) pandemic created a situation of severe uncertainty fuelled by disruptions finance, politics, social life and healthcare domains. As such, the pandemic constitutes an immense psychological challenge and puts at risk people’s mental health.

First studies have reported adverse psychological consequences of the pandemic among people with and without pre-existing mental health conditions. Patients with anxiety, depression, bipolar disorders, schizophrenia (3–5) and obsessive-compulsive disorder (OCD) (6) were reported to experience a pandemic-related increase in symptoms. Likewise, the general public reported worsened mental health with a rise primarily seen in anxiety and depression levels (7–12).

OCD is likely to be disproportionally affected by the pandemic. Many OCD symptoms evolve around contamination and cleaning (13), the key focus of Covid-related information campaigns. These news coverages and governmental guidelines may thus give validity to OCD-symptomatic behaviour, potentially intensifying or triggering them. Indeed, first evidence suggested a worsening of symptoms in OCD patients during the pandemic (6). Moreover, as contamination-related concerns and behaviours increase in the general public, obsessive-compulsive (OC) symptoms may also be triggered in non-patient populations.

Elevated levels of psychiatric symptoms after a stressor, such as the pandemic onset, are common. Decades of research demonstrated that mental health worsens after stressful events (14,15). However, importantly, psychiatric levels usually improve after a reasonable time without inflicting long-term impairments (16–18). This adaptation process has been attributed to coping and (re-)appraisal strategies (19). A failure of this process, however, may lead to long-term adverse consequences and chronic mental-health problems (16,17). It is thus critical to not only assess momentary increases in psychiatric symptoms during the pandemic, but to also investigate the long-term trajectories of these symptoms.

Here, we conducted a large-scale longitudinal study investigating the impact of the Covid-19 pandemic on OC, anxiety, and depression symptoms. We tracked these psychiatric scores over a period of several months and show that OC symptoms continuously increase, whilst anxiety and depression level and diminish. Moreover, we investigated how these symptoms impacted Covid-related information seeking and adherence to governmental guidelines, based on the hypothesis that OCD drives excessive information seeking (20–24). We show that rising OC symptoms drive pandemic-related information seeking which in turn promotes higher guideline adherence.

## Materials and methods

### Study design and procedure

We conducted a longitudinal online study of the general public, collecting data at two time points throughout the first pandemic wave (May - August 2020; Figure 1). At both time points, participants reported OC (25), anxiety and depression symptoms (26) using standardised questionnaires. We also recorded participants’ Covid-19-related information seeking with a new scale. At the first time point (T1), participants also indicated their baseline news and social media consumption and completed a cognitive ability assessment (27). At the second time point (T2), we additionally measured people’s adherence to Covid-19 guidelines as instructed by the UK government.

**Figure 1.**
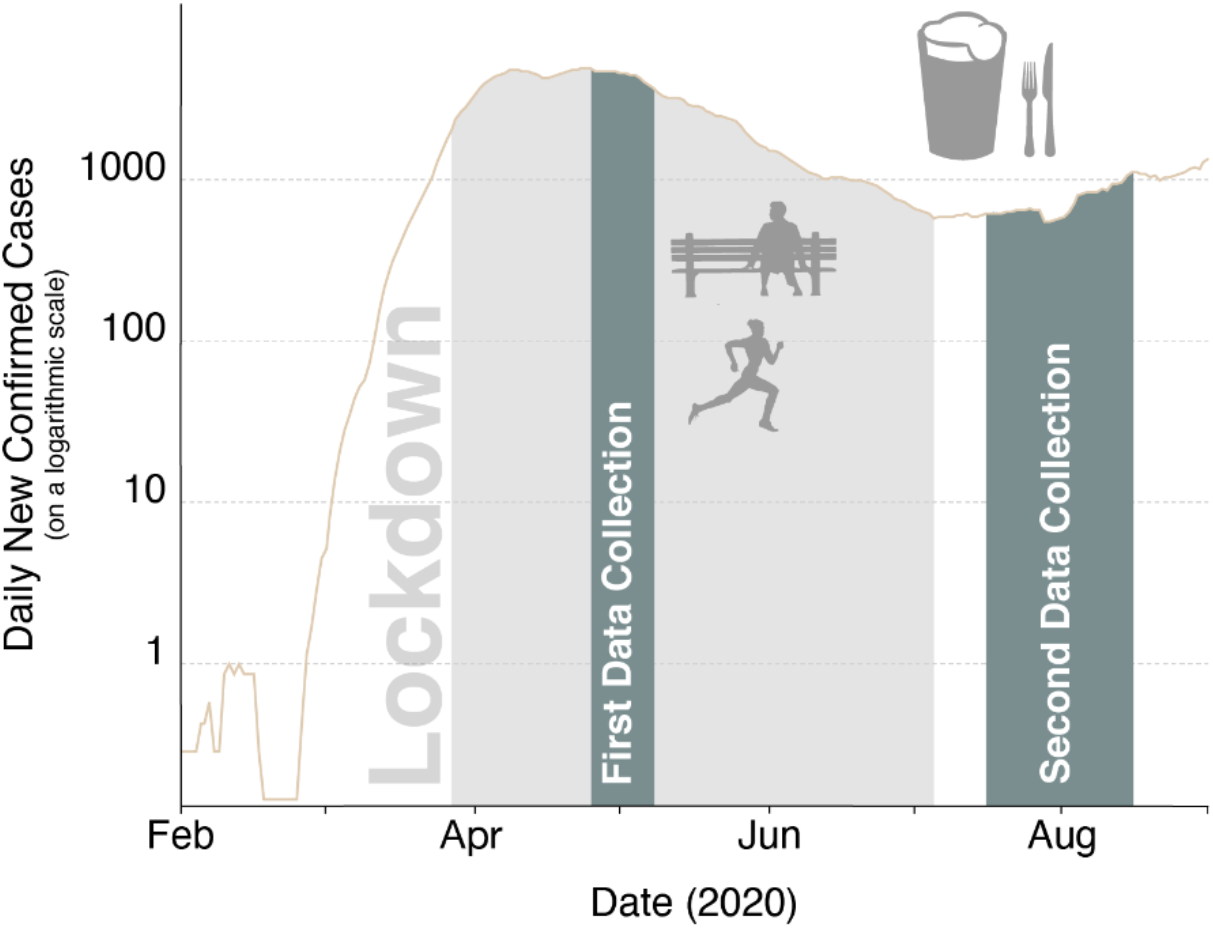
Longitudinal data collection of mental health symptoms. The first data collection took place from 24^th^ of April to 7^th^ of May. Shown is the log-average of daily new confirmed Covid-19 cases, using the rolling 7- day average in the UK from 1^st^ of February 2020 to 1^st^ of September 2020. Lockdown was formally enforced on 26^th^ of March. The ease of lockdown commenced on the 10^th^ of May when people were encouraged to visit parks again and engage in unlimited outdoor exercise. On the 4^th^ of July pubs and restaurants were among the last businesses to come out of lockdown. The second data collection took place from the 15^th^ of July until the 15^th^ of August, after the lockdown restrictions have been lifted. The pandemic curve has been plotted on the basis of data maintained by Our World in Data(28).

Data collection for T1 lasted from the 24th of April 2020 (first pilot) until the 7th of May 2020. This was approximately at the peak of the first pandemic wave in the UK (Figure 1). The second data collection (T2) directly followed the largest lift in pandemic-related restrictions in the UK (i.e. reopening of restaurants and pubs) which allowed us to assess the adaptation of mental health symptoms to a substantial environmental change. The T2 data for was collected between the 15^th^ of July 2020 and the 15^th^ of August.

### Participants

We recruited participants living in the UK via the Prolific recruiting service (https://www.prolific.co/). All subjects were over 18 and gave informed consent before starting the study. The study was approved by University College London’s Research Ethics Committee.

A total of 446 participants completed the study at T1. We excluded 20 participants because of missing data or failing at least one of two attention checks (instructed questionnaire answers). This resulted in a final T1 sample of 416 participants (237 females; *M*_*age*_=34.0, *SD*_*age*_= 12.536).

We invited this final T1 sample to take part at T2. Of the re-invited participants, 325 completed the second time point of which we excluded additional 21 participants because of missing data or failed attention checks. The final T2 sample consisted of 304 participants (167 females; *M*_*age*_=35.0, *SD*_*age*_=12.565). This led to an overall retention rate of 78%. We did not observe any differences in mental health symptoms (at T1) between subjects that did versus did not complete the T2 study as indicated by Welch’s two sample t-tests (OC symptoms: *t*(180.13)=1.662, *p*=0.098; anxiety: *t*(212.33)=0.769, *p*=0.443; depression: *t*(221.13)=0.359, *p*=0.720).

### Questionnaires

We implemented all questionnaires using a web API programmed with React JS libraries (https://reactjs.org/).

#### Standardized questionnaires

To assess the impact of the Covid-19 pandemic on OC, anxiety and depression symptoms, we administered self-report psychiatric questionnaires. We assessed OC symptoms using the Padua Inventory-Washington State University Revision (PI-WSUR) (25). We chose the PI-WSUR because of its good test-retest reliability (29) and detailed sub-scales. These subscales enabled us to look at changes in scores over repeated testing and assess sub-scores of interest, such as the contamination obsessions and washing compulsions sub-scores (COWC).

We further measured anxiety and depression, two psychiatric domains that are often comorbid (13) with OCD and show a substantial overlap in the general population, using the Hospital Anxiety and Depression Scale (HADS) (26). The HADS consists of two sub-scales (anxiety, depression) that are intended to be evaluated separately. We chose the HADS because of its high internal consistency and test-retest reliability (30).

#### Development and validation of the Covid-19 related information-seeking questionnaire

To investigate information-seeking during the pandemic and across different psychiatric domains, we developed a new Covid-19 information-seeking questionnaire. The questionnaire entailed 5-items with a 5-point Likert-scale with lower scores indicating less information seeking. Items asked about information exchange and seeking via different social (and) media channels (Supplemental Table 1). We assessed the dimensionality of our questionnaire using a principal component analysis (PCA; cf. Supplemental Information), internal consistency examining Cronbach’s Alpha coefficient and test-retest reliability looking at Pearson’s correlation of T1 and T2 scores (cf. Supplemental Information).

#### Baseline news and social media consumption

To control for potential confounds unrelated to the pandemic, we also asked participants to retrospectively indicate their average weekly news and social media consumption before the onset of the pandemic (i.e. November 2019; Supplemental Table 3). To ensure the robustness of our main findings, we repeated regression models assessing information seeking with baseline news consumption and social media score as a covariate.

#### Adherence to governmental guidelines

To investigate concrete behavioural links of information seeking and psychiatric scores, we asked participants to indicate to which degree they followed recommendations from authorities (Supplemental Table 4). We administered these questions after the ease of lockdown at T2 as by then there were substantial behavioural guidelines put in place by the UK government to prevent the spread of Covid-19. In the rest of the paper, we will refer to the total score as *guideline adherence* score.

### Statistical analyses

We pre-processed and analysed data in MATLAB 2020a (MathWorks), and additionally used SPSS Statistics (version 26, IBM) and R version 1.2.5033 (2019) for further data analysis.

To investigate whether psychiatric scores changed differently over time, we conducted a repeated-measures ANOVA with two within-subject factors (F1: psychiatric domain, F2: time point) and Huynd-Feldt corrections. For comparability between scores, we first z-scored all scores across both time points. To further compare the changes of psychiatric scores and information seeking between time points, we conducted paired t-tests using the *stats* package in R, supplemented with paired permutation tests using the *CarletonStats* package(31,32).

To evaluate the association of Covid-19 related information seeking and psychiatric scores for each time point separately, we computed robust multiple regression models using the *rlm* of the *MASS* package in R with a bisquare method to down-weight outliers (33). To investigate changes in the association between information seeking and psychiatric scores over time, we also conducted a random intercept mixed-effects model using the *lm4* package (34). The model entailed main effects for time point (coded as 0 (*T1*) and 1 (*T2*)) and all z-scored psychiatric scores as well as interactions between time point and psychiatric scores. In the syntax of the *lme4* package, the model was specified as follows:

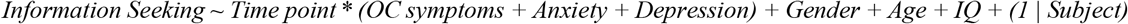

Finally, we conducted a mediation analysis to examine whether information seeking was associated with guideline adherence. For this analysis we used the Mediation Toolbox in MATLAB (https://github.com/canlab/MediationToolbox) which adapts the standard 3-variable path model (35,36). All results refer to two-tailed bootstrapped p-values and the procedure was repeated 10,000 times with replacement producing a null hypothesis distribution. In our mediation model, the total score on OC symptoms at T1 was taken as a predictor, guideline adherence at T2 as the dependent variable and information seeking at T1 as the mediator (cf. Supplemental Information for control adaptations of the hypothesized mediation model).

All regression models covaried for age, gender and IQ and we z-scored all included variables except for gender (for better interpretability) prior to the analysis to allow comparability of regression coefficients. To ensure that multicollinearity was not driving any of our results, we examined the variance inflation factor (VIF) for all multiple regression models. The VIF was below 3 for all models and thus below the standard cut-off score of 10 (37).

## Results

To investigate the impact of the Covid-19 pandemic on OC, anxiety, and depression symptoms, we tested a general-public UK sample longitudinally at two time points during and after the first pandemic wave.

### Heightened psychiatric symptom scores during the Covid-19 pandemic

We tested the hypothesis that the Covid-19 pandemic was associated with an overall increase in psychiatric symptom scores that have been linked to OCD. As expected, OC symptoms, anxiety and depression scores were all elevated. To assess OC symptoms, we used the PI-WSUR total score, which, with a mean of 31.643 (*SD*=23.026; cf. Figure 2A) at T1, showed clearly elevated levels compared to previous population samples (25,38–40). Some sub-scores revealed scores similar to OCD patient samples (Supplemental Figure 2) (25). Our findings thus strongly suggest that OC symptoms were substantially elevated at the peak of the first pandemic wave.

**Figure 2.**
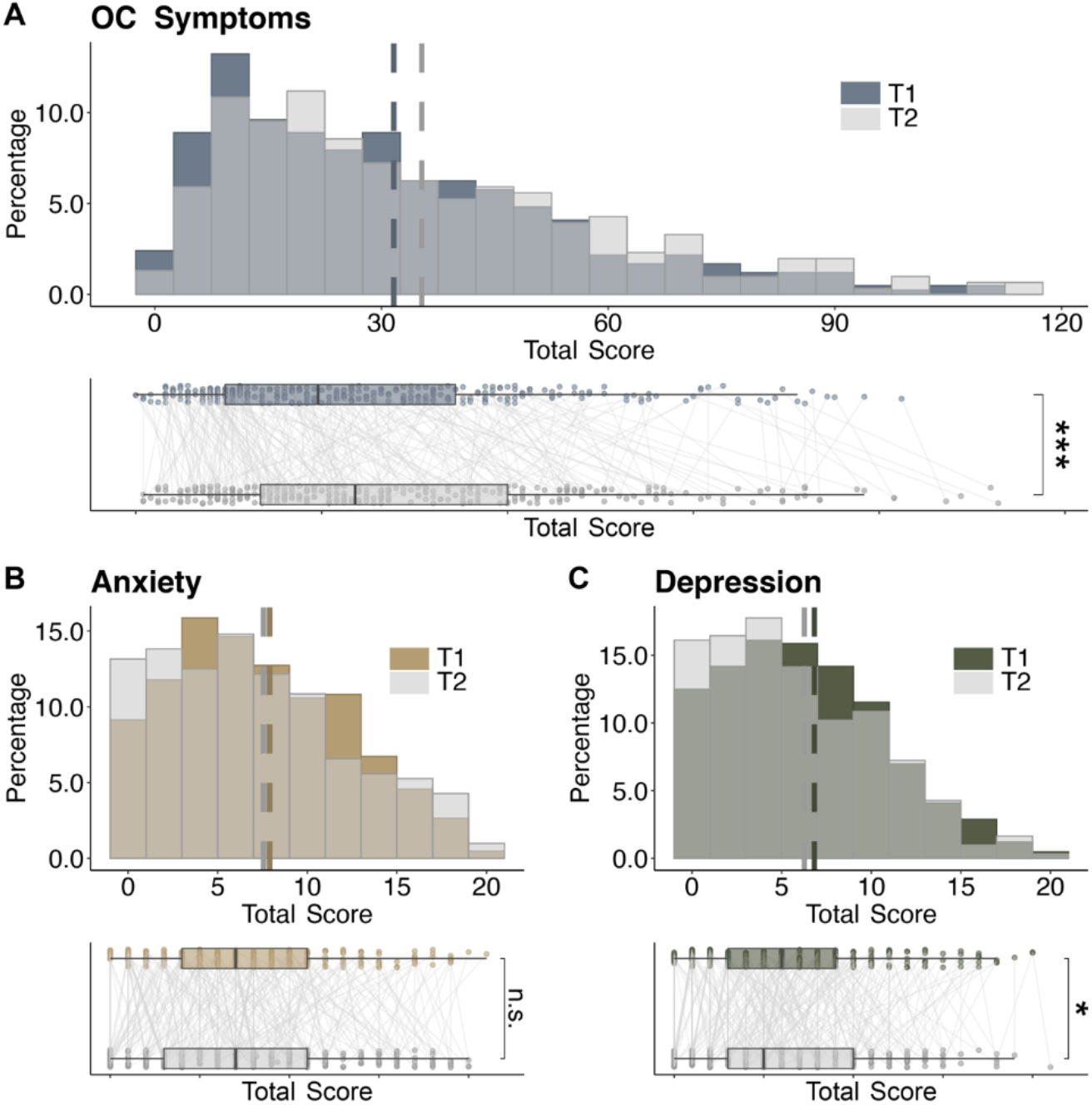
Changes of psychiatric symptoms during the first Covid-19 pandemic wave. Top-panels show the distributions of OC symptoms **(A)** as measured by PI-WSUR, anxiety (**B**) and depression scores (**C**) of the Hospital Anxiety and Depression Scale (HADS) at time point 1 (T1; N=416) and time point 2 (T2; N=304). Dashed lines denote the means of each time point. Bottom panels show boxplots for each of the psychiatric symptoms. Boxplots visualize an increase in OC symptom scores from T1 to T2 (A), stable levels of anxiety scores (B) and a decrease in depression scores (C). Connected thin lines show individual scores for T1 and T2 (N=304). * - *p*< 0.05, *** - *p*< 0.001, n.s. – non-significant.

Similarly, we found heightened anxiety and depression scores. Almost half of the participants reached clinical cut-off scores. At T1, 48.56 % (202/412) of all participants scored above the standardised cut-off score of 8 on anxiety (*M*=6.825, *SD*=4.471; cf. Figure 2B) and 41.35% (172/412) scored above the cut-off score of 8 on depression (*M*=7.898, *SD*=4.840; cf. Figure 2C). In comparison, one of the largest non-patient study administering the HADS (*N=*2481) prior to Covid-19 has recorded a rate of 21% and 23% above the cut-off score for anxiety and depression respectively (41). Overall, our results show a systematic and consistent elevation across all assessed psychiatric symptom scores in the general public at the peak of the first pandemic wave.

#### Only OC symptoms rise further during the pandemic

We next assessed the trajectories of these psychiatric symptoms throughout the pandemic. Leveraging the longitudinal design of our study, we assessed whether symptoms showed signs of adaptation, a i.e. decrease, or whether they rose further(19,42).

To do so, we ran a 2-way repeated measures ANOVA with within-subject factors time (T1 vs. T2) and psychiatric domain (OC, anxiety, and depression symptoms). We found a significant time by psychiatric domain interaction (*F*(1.976, 592.717)=19.933, *p*<0.001) extending a significant main effect of time (*F*(1, 300.000)=4.393, *p=*0.037).

To further inspect these effects, we compared T1 and T2 scores for each psychiatric domain separately. Importantly, we found that OC symptoms further increased during lockdown with higher average scores at T2 compared to T1 (*t*(303)=6.389, *p*<0.001; Figure 2A; cf. Supplemental Information and Supplemental Figure 3 for non-parametric tests). Next, we tested whether this also held true when ignoring the symptoms most closely linked to the pandemic, by excluding items from the contamination and washing (COWC) subscale. Interestingly, we found the same increase in OC symptoms without COWC sub-scores (*t*(303)=4.983, *p*<0.001), suggesting that OC symptoms throughout the lockdown did not show an adaptive decrease, but rather a worrying escalation.

This increase in symptoms was unique to the OC domain. In fact, we saw a significant decrease in depression scores from T1 to T2 (*t*(303)=-1.985, *p=* 0.048; Figure 2C). When analysing anxiety symptoms, we did not observe a significant change between T1 and T2 (*t*(303)=-0.266, *p*=0.791; cf. Figure 2B). These findings thus suggest that whilst depression and anxiety symptoms adaptively decreased or remained on the same level, OC symptoms further rose throughout the lockdown.

### Psychiatric dimension scores and information-seeking behaviour

Next, we assessed whether the psychiatric symptom scores were driving pandemic-related behaviours. In particular, given the prior account of increased information seeking in people with high OC symptoms (20–24), we expected individuals to engage in more Covid-related information seeking.

#### Information seeking over the course of the Covid-19 pandemic

We thus examined whether people engaged in Covid-related information seeking using a newly developed and validated scale (cf. Supplemental Information). We assessed information seeking at both time points, allowing us to trace how this behaviour changed throughout the course of the pandemic. We found that information seeking decreased from T1 to T2 (*M*_*T1*_= 17.018; *SD*_*T1*_= 4.194, *M*_T2_= 14.625, *SD*_T2_= 4.317, *t*(303)=-9.951, *p*<0.001; Figure 3 and Supplemental Figure 5). This means that participants engaged in Covid-related information seeking, and did so primarily at the beginning of lockdown, when little was known about the pandemic.

**Figure 3.**
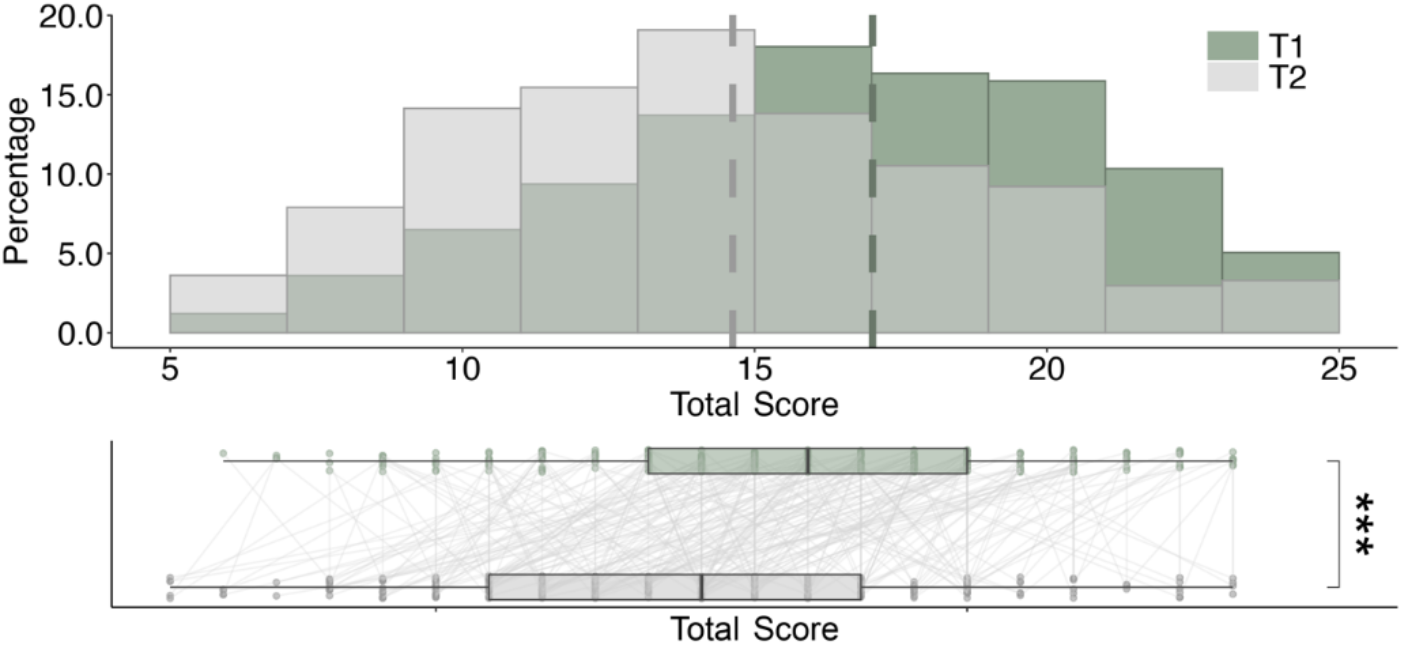
Histogram of the Covid-19 related information seeking scores at time point 1 (T1; N=416) and time point 2 (T2; *N*=304). Dashed lines denote the means of the corresponding distributions. The lower panel shows boxplots constructed with the paired data available at both time points (*N*=304). Thin lines connect individual scores between the two time points. ***- *p*< 0.001.

#### OC symptoms were uniquely associated with increased pandemic-related information seeking

We next investigated whether information seeking was associated with any of the psychiatric symptom scales. We found that OC symptoms indeed were linked to increased information-seeking at both time points (*r*_*T1*_(414)=0.218, *p*<0.001, *r*_*T2*_(302)=0.222, *p*<0.001). A similar, but smaller association was found for anxiety scores (*r*_*T1*_(414)=0.180, *p*<0.001, *r*_*T2*_(302)= 0.145, *p*=0.012). We did not find a significant association between depression and information seeking (*r*_*T1*_(414)=0.096, *p*=0.051, *r*_*T2*_(302)= 0.077, *p*= 0.182). These results held when controlling for demographic variables such as age, gender and IQ (cf. Figure 4; T1: OC symptoms_T1_*β*=0.230, *p*<0.001; anxiety_T1_ *β*=0.174, *p=* 0.001; depression_T1_ *β*=0.084, *p*= 0.106; T2: OC symptoms_T2_ *β*=0.230, *p*<0.001; anxiety_T2_ *β*=0.154, *p=*0.013; depression_T2_ *β*=0.08, *p*= 0.164). Moreover, these results also replicated when additionally controlling for Covid-unrelated news and social media consumption (retrospective baseline measure; OC symptoms *β*_*T1*_=0.214, *p*<0.001, *β*_*T2*_=0.206, p=0.001; anxiety *β*_*T1*_=0.181, *p*<0.001, *β*_*T2*_=0.140, *p*=0.018).

**Figure 4.**
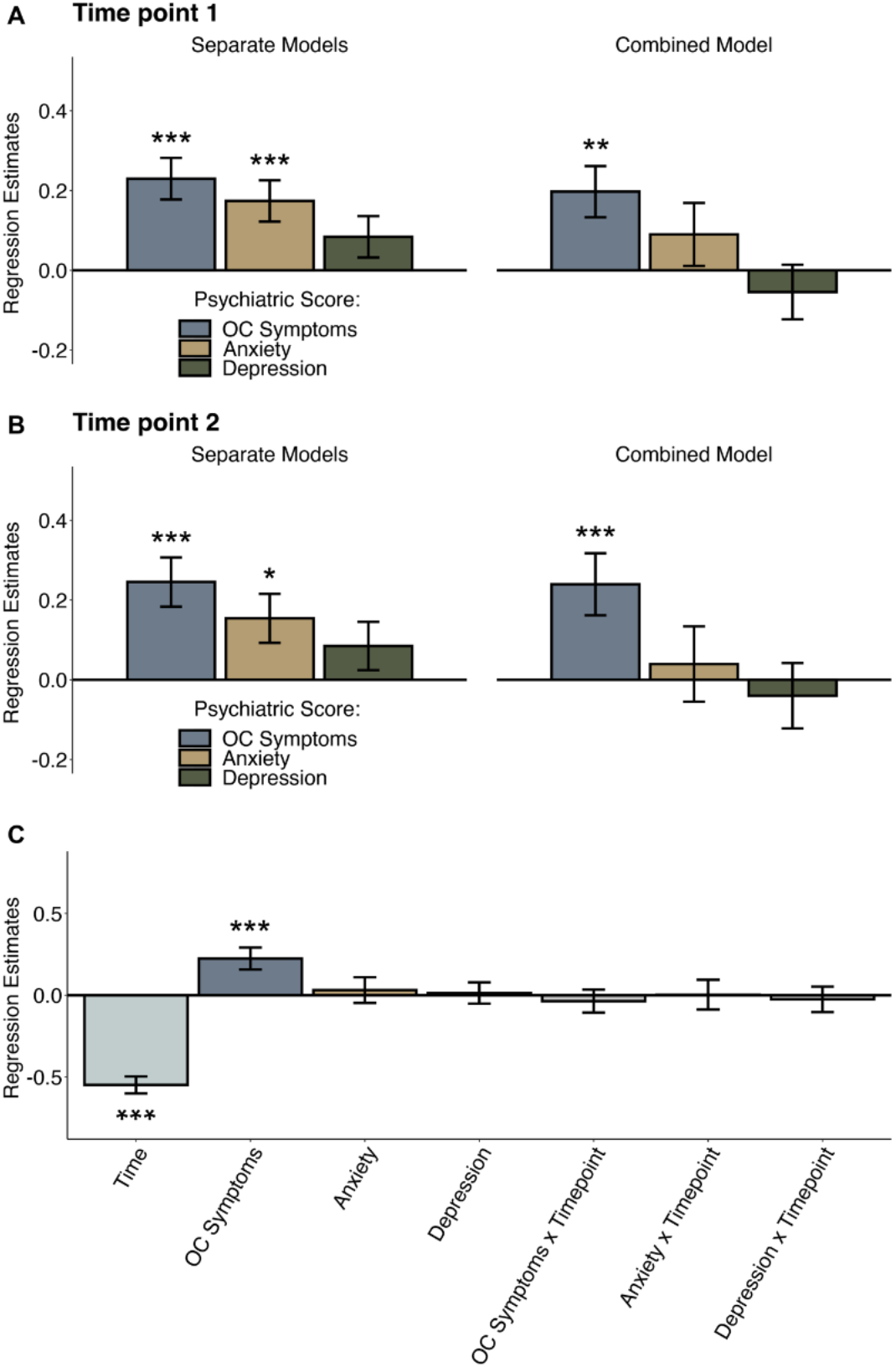
Regression analysis showing the association between Covid-19 related information seeking and psychiatric scores (OC symptoms, anxiety and depression). **(A-B)** Regression models estimated separately for each psychiatric dimension (left panels) showed that OC symptoms as well as anxiety were associated with increased information seeking at time point T1 (*N*=416; A) and time point T2 (*N*=304; B). When combining all three psychiatric scores in one model (right panels) only OC symptoms remained associated with information seeking at both time points. Estimates from the mixed-effects regression model assessing the effect of psychiatric scores, time point and their interactions on information-seeking showed that time point was negatively associated with information seeking (*N*=304; **C**). This indicates that information seeking significantly decreased from T1 to T2, and OC symptoms remained positively associated with information seeking. None of the remaining main and interaction effects (OC symptoms x Time point, Anxiety x Time point, Depression x Time point) were significant. Error bars represent standard errors and * - *p*<0.05, ** - *p*<0.01, *** - *p*<0.001.

These consistent correlations between psychiatric symptoms and information seeking were not surprising considering the psychiatric domains themselves are known to be related and are thus expected to show similar links (correlations between symptoms scores in this sample: OC symptoms and anxiety: *r*_T1_(414)= 0.598, *p*<0.001, *r*_T2_(302)= 0.631, *p*<0.001; OC symptoms and depression: *r*_T1_(414)=0.385, *p*<0.001, *r*_T2_(302)=0.455, *p*<0.001; anxiety and depression: *r*_T1_(414)=0.675, *p*<0.001; *r*_T2_(302)=0.687, *p*<0.001; cf. Supplemental Figure 1).

To assess which of the psychiatric domains uniquely explained increased Covid-related information seeking, we conducted a regression including all psychiatric symptoms as predictors of information seeking. This analysis revealed that only OC symptoms were uniquely associated with information seeking (*β*_*T1*_=0.197, *p*=0.002; Figure 4), and that there was no longer any association with anxiety (*β*_*T1*_=0.089, *p*=0.256). The same effects were found when investigating T2 scores (OC symptoms: *β*_*T2*_=0.240, *p=*0.002; anxiety: *β*_*T2*_=0.04, *p*=0.675). Again, including baseline news and social media consumption as covariates did not change the results (OC symptoms: *β*_*T1*_=0.165, *p*=0.008, *β*_*T2*_=0.186, *p=*0.014; anxiety: *β*_*T1*_=0.092, *p*=0.229, *β*_*T2*_=0.028, *p*=0.754).

Finally, we investigated how changes in psychiatric scores related to changes in information seeking. We constructed a mixed-effects regression model with information seeking as the dependent variable and time point, psychiatric symptoms, as well as their interactions, as predictors (see Methods). Time point was negatively associated with information seeking (*β*=-0.548, *SE* = 0.052, *p*<0.001) confirming the observed decrease in information seeking from T1 to T2 (Figure 4C). We further found a main effect of OC symptoms showing that they were linked to higher Covid-related information seeking (*β*=0.225, *SE* = 0.067, *p*= 0.001). No significant interaction effects were found between the psychiatric factors and time point (time point*OC symptoms: *β*=- 0.036, *p*=0.608, time point*anxiety: *β*=0.003, *p*=0.608, time point*depression: *β*=-0.025, *p*=0.748). These results are in support of OC symptoms as the main driving factor for information-seeking related to the pandemic.

#### OC driven information seeking promotes guideline adherence

To investigate whether and how Covid-related information seeking translated into practically relevant behaviours we also measured participants’ adherence to governmental guidelines aiming to control the pandemic once the lockdown was eased (cf. Methods for details). We found that information seeking was indeed directly linked to increased guideline adherence (*r*(302)=0.151, *p*=0.008). Similarly, we found that OC symptoms predicted increased guideline adherence (*r*(302)=0.117, *p*=0.042). Thus, not only were OC symptoms and increased information seeking linked, but both variables also showed a positive relationship with the adherence to government-imposed Covid-guidelines.

To closer investigate this relationship we performed a mediation analysis with all three factors (i.e. OC symptoms, information seeking and guideline adherence). The mediation analysis showed a significant direct effect of OC symptoms at T1 on guideline adherence at T2 (*β*=0.14, *p=*0.024, path *c*) and on information seeking at T1 (*β*=0.23, *p*<0.001, path *a*). It further showed a significant path from information seeking at T1 to guideline adherence at T2 (*β*=0.13, *p=*0.025, path *b;* see Figure 5). Thus, OC symptoms were associated with information seeking as well as guideline adherence, and information seeking and guideline adherence themselves were associated too. Importantly, we found a significant mediation effect via information seeking (*β*=0.03, *p*=0.018, path *ab*). When then controlling for information seeking, the effect of OC symptoms on guideline adherence was no longer significant (*β*=0.11, *p*=0.091, path *c’*). These results suggest that individuals scoring higher on OC symptoms engaged more in pandemic-related information seeking, which in turn reinforced their adherence to governmental guidelines (cf. Supplemental Information for control adaptations of this mediation model).

**Figure 5.**
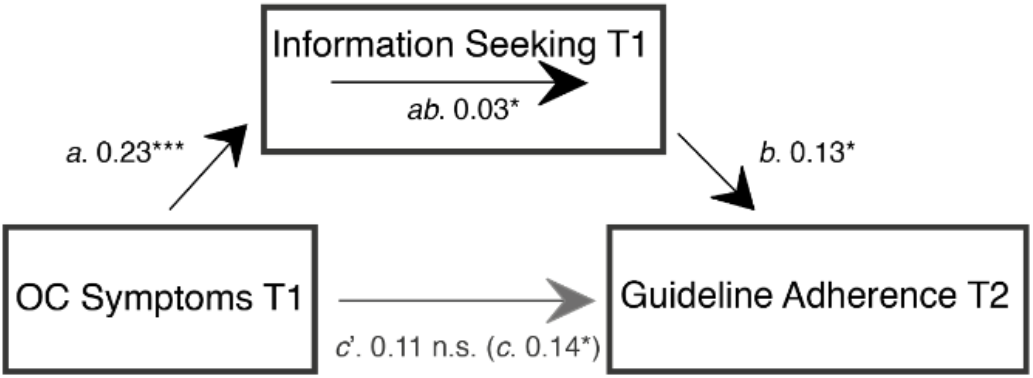
Mediation model assessing the relationship between OC symptoms and information seeking at T1 and guideline adherence at T2. Effect of OC symptoms on guideline adherence was mediated by information seeking. Displayed are standardized regression coefficients. Mediation analysis was performed on data from participants who completed the study at both time points (*N*=304) *- *p<*0.05, *** - *p*<0.001. * - n.s. – non-significant.

## Discussion

Understanding how psychiatric symptoms change throughout the Covid-19 pandemic is essential when making inferences about the crisis’ long-term mental-health consequences. We studied multiple psychiatric domains longitudinally during the first pandemic wave and show an elevation across all. However, while one of the psychiatric symptoms decreased another remained stable over time, we found a further rise of OC symptoms even though the peak of the first pandemic wave had passed. Our findings highlight that OC symptoms are disproportionally affected by the pandemic by documenting their selective increase throughout the pandemic for the first time, which may result in serious adverse long-term consequences.

The here documented elevations of anxiety and depression scores during the pandemic are in line with previous studies conducted of the (UK) general public (7–12). For the first time, we also report a heightening in OC symptoms in the general public with levels exceeding previous non-patient populations, in part matching patient levels (25,38–40). This observation is of particular importance as OCD is likely to be disproportionally affected by this pandemic with many of the governmental guidelines directly promoting OCD symptoms (43). Furthermore, OC symptoms were also systematically elevated in domains not directly linked to Covid-related themes (e.g. increased checking vs. obsessional thoughts about harm to self or others and contamination obsessions and washing compulsions) speaking for a generalized and thus even more worrying effect.

In contrast to previous studies that looked at the psychiatric symptoms (before and) at one time point during the pandemic (8,9), we used a longitudinal design that allowed us to characterize the dynamics of psychiatric symptoms throughout the pandemic. Based on coping and stress adaptation theories (19,42) we expected a decrease in symptoms after the peak of the pandemic which has been recently documented for some psychiatric domains (11). However, we found an important dissociation in symptom dynamics. While depression decreased and anxiety levels plateaued with the ease of lockdown and the drop of Covid-19 infections, the OC symptom levels further increased. This increase spanned several sub-domains which may indicate that it was not purely elicited by health worries but uncertainty and stress during the pandemic. These prolonged high levels of OC symptoms are worrying and might develop into a disorder. It is thus critical to continue to closely observe their development.

We further investigated how OC symptoms related to daily behaviours such as information seeking and adherence to governmental Covid-guidelines. Previously, increased information seeking has been reported in subjects with high OC symptoms in abstract laboratory tasks (20–24). Improving ecological validity, we developed a novel information-seeking measure related to Covid-19. We indeed found that individuals with high OC scores tended to seek more information, even when controlling for other psychiatric symptoms, the general news and media consumption, and demographic variables. Furthermore, this association between OC symptoms and information seeking persisted throughout the pandemic.

We further show that Covid-related information seeking was critical for later adherence to governmental guidelines. We found that information seeking mediated the influence of OC symptoms on guideline adherences, meaning that individuals with elevated OC symptoms stronger adhered to guidelines as they extensively sought Covid-related information. Our findings thus suggest that OC symptoms, albeit burdening for the individual, can be beneficial to society as they promote efforts to contain the virus.

Our study has some unique strengths such as its large sample size, longitudinal testing and real-world relevance of the measures. However, it relies on self-reported measures which might not accurately reflect actual behaviours and their changes. It would be interesting to further investigate these associations using additional metrics, such as smartphone geolocation patterns, browser history, or in-lab experiments. Moreover, even though our study exploits its longitudinal design to report directional effects, causality cannot directly be inferred.

Overall, this study provides a first account of OC-symptom dynamics in the general public throughout Covid-19 pandemic and how these symptoms translate into other behaviours such as information seeking and adherence to governmental guidelines. The increase and persistence of OC symptoms should serve as a warning sign stressing the importance of close monitoring of individuals at risk of developing OCD. It calls for treatment interventions and public mental-health support to prevent long-term psychiatric problems.

## Data Availability

Fully anonymised data for this study will be available from the corresponding authors upon reasonable request with peer-reviewed publication. Simultaneously, code for data analysis will be available from a dedicated Github repository (https://github.com/DevComPsy).

## Acknowledgements

A.M.L. is a pre-doctoral fellow of the International Max Planck Research School on Computational Methods in Psychiatry and Ageing Research (IMPRS COMP2PSYCH). The participating institutions are the Max Planck Institute for Human Development, Berlin, Germany, and University College London, London, UK. For more information, see: https://www.mps-ucl-centre.mpg.de/en/comp2psych. T.U.H. is supported by a Sir Henry Dale Fellowship (211155/Z/18/Z) from Wellcome & Royal Society, a grant from the Jacobs Foundation (2017-1261- 04), the Medical Research Foundation, and a 2018 NARSAD Young Investigator grant (27023) from the Brain & Behavior Research Foundation. T.U.H. is also supported by the European Research Council (ERC) under the European Union’s Horizon 2020 research and innovation programme (grant agreement No 946055). V.S. received funding from the European Union’s Horizon 2020 research and innovation programme under the Marie Skłodowska-Curie grant agreement No 895213.

## Supplemental Information

### Psychiatric symptom scores

Previous research has shown that OCD, anxiety and depression are highly comorbid and are usually correlated in dimensional samples (13,44,45). We replicated these findings in our sample (Supplemental Figure 1).

**Supplemental Figure 1.**
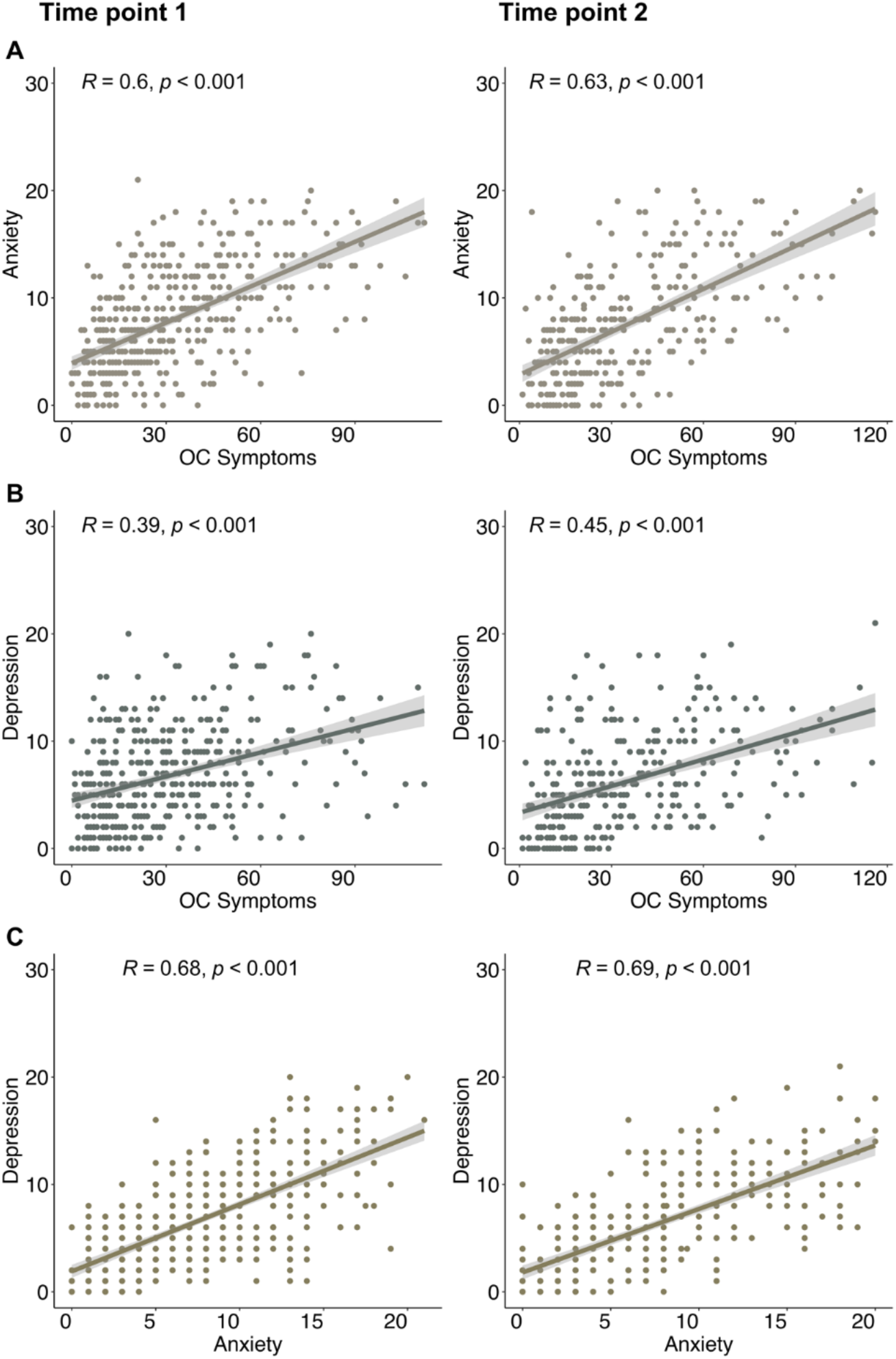
Psychiatric symptom score associations at both time points. Pearson’s pair-wise correlations between psychiatric scores (OC symptoms, anxiety and depression; **A-C**) at T1 **(left panel)** and T2 **(right panel)**. Shaded areas represent 95% confidence intervals.

### OC symptom sub-scores: Our sample versus samples as reported by previous literature

To put the OC symptom scores from the PI-WSUR measured in our sample (*N*_*T1*_=416; *N*_*T2*_=304) into perspective, we compared mean scores to scores reported in the literature (25,38–40). We restrained from quantitative comparisons due to unequal sample sizes and other potential confounding factors (e.g. age). On average, our sample scored similarly high on the washing and contamination sub-scale (COWC) than the patient samples of Burns et al. (*N*=15) and an unpublished patient sample of Hauser et al. (*N*=31) at both time points(25). On all other sub-scales, our sample scored somewhat lower than the patient samples, but clearly higher than two non-patient population samples (collected before the pandemic). Only for ‘harm to self or others’ (OITHSO) and ‘dressing and grooming compulsions’ (DRGRC), the non-patient sample reported by Burns et al. scored similarly(25). Thus, overall our sample seems to score between patient and non-patient samples with clearly elevated scores in most sub-domains.

**Supplemental Figure 2.**
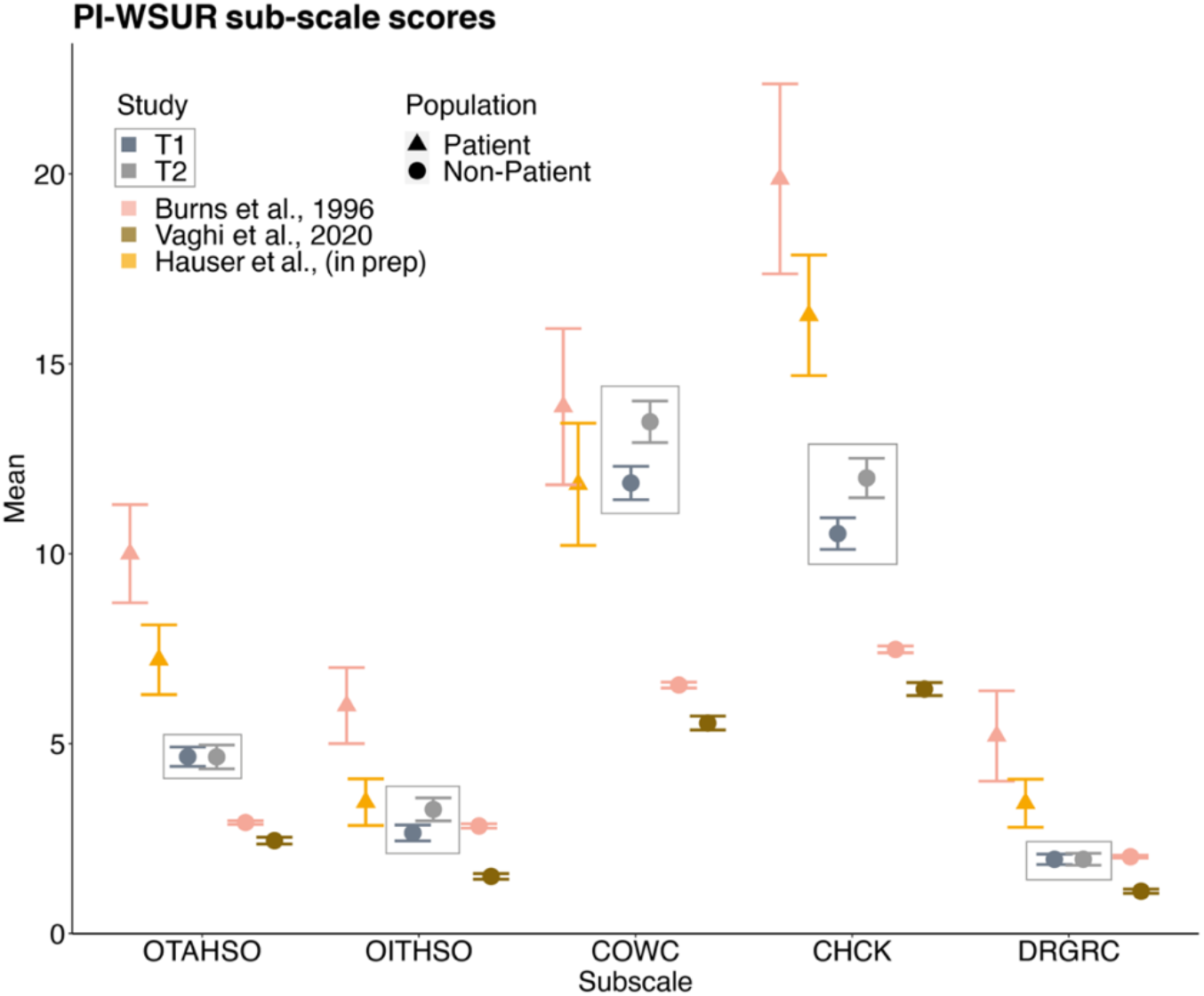
Average sub-scale scores of the Padua Inventory Washington State University Revision (PI-WSUR) measuring OC symptom domains in T1 and T2 in comparison to previous studies. Current study summary statistics are highlighted by a surrounding square (*N*_T1_=416; *N*_T2_=304). These are contrasted with two patient samples reported by Burns et al. (*N*=15) displayed as pink triangles and unpublished patient-data by Hauser et al. (*N*=31), displayed as yellow triangles(25). A non-patient sample reported by Burns et al. (*N*=5010) as pink dots and data from a previously collected young UK public sample reported by Vaghi et al. (*N*=1606) as brown dots(25,38). The comparison shows that on average our sample scored in-between the patient and non-patient samples reported in the literature indicating a heightened OC symptom level across most sub-domains. Abbreviations: OTAHSO- Obsessional Thoughts about Harm to Self or Others, OITHSO- Obsessional Impulses about Harm to Self or Others, CHCK- Checking Compulsions, COWC- Contamination Obsessions and Washing Compulsions, DRGRC Dressing and Grooming Compulsions. Data points are means, and error bars represent standard errors.

### Differential changes in psychiatric scores throughout the pandemic

To evaluate the robustness of our evidence for differential changes in psychiatric dimensions, we additionally performed permutation tests to compare the observed differences in means 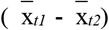 to a reference null distribution (Supplemental Figure 3). The observed difference in mean OC symptoms of T1 and T2 was significantly greater than the null distribution (*p*<0.001), confirming the increase in OC symptom scores between the two time points. This result held true when excluding the contamination and washing sub-scores (*p*<0.001). In contrast, the difference in mean depression was significantly smaller than the null distribution (*p*= 0.047), confirming a decrease in depression scores. The observed difference in mean anxiety was not significantly different from the null distribution (*p*=0.796), thus the permutation test also did not support a change in anxiety scores from T1 to T2. These results indicate that already heightened OC symptom scores further increased from mid-way through the first pandemic wave to after it, in the absence of any changes in mean anxiety scores and a contrasting decrease in mean depression scores.

**Supplemental Figure 3.**
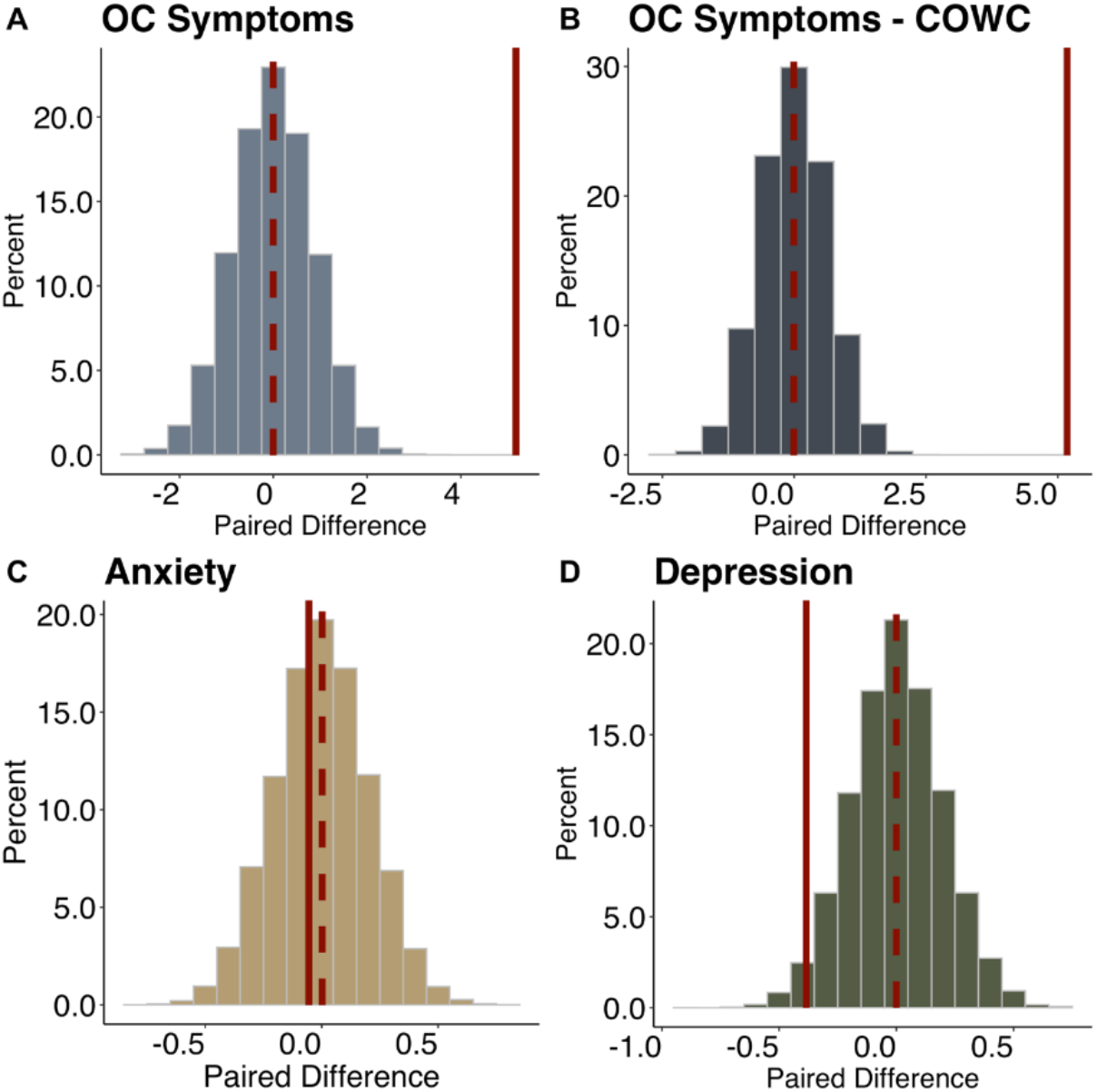
Histograms of the null distributions of the mean differences in psychiatric scores between T1 and T2 (*N*=20,000 permutations) for OC symptoms (A), OC symptoms scores without the contamination obsessions and washing compulsions (COWC) sub-scores (B) and anxiety (C) and depression symptoms (D). The solid red lines represent the observed difference in means while the dashed red line represents the mean difference observed for the shuffled data. The OC symptoms – COWC scores have been computed by excluding the COWC sub-score from the PI-WSUR total score for each subject. OC symptom scores with and without COWC, as well as depression scores were more extreme than the shuffled data (Both OC scores: *p*<0.00; depression: *p*= 0.047).

### A novel Covid-19-related information-seeking questionnaire

We developed a new questionnaire measuring Covid-19 related information seeking. The questionnaire measured news and statistics consumption via different media channels and the general information exchange with other individuals. The items are displayed below in Supplemental Table 1.

**Supplemental Table 1.**
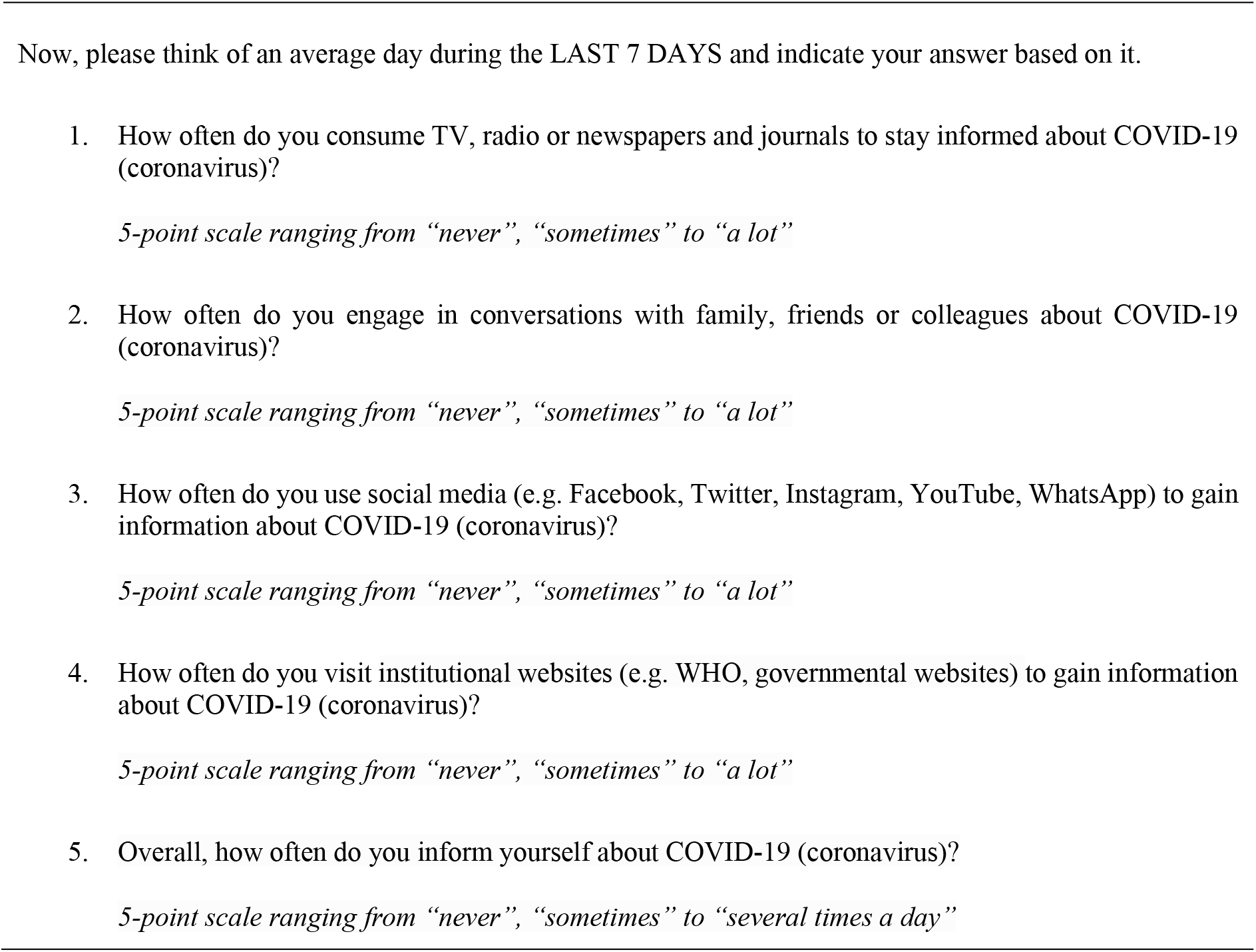
Items of the Covid-19-related information-seeking questionnaire.

### Validity and psychometric properties of the new pandemic-related information-seeking questionnaire

To assess the validity of our Covid-19 information-seeking questionnaire we ran a PCA analysis to investigate its dimensionality. For this analysis, we used the data of all participants at T1 that had completed the information-seeking questionnaire and had not failed any attention checks [*N*=420 (235 females; *M*_*age*_=34, *SD*_*age*_= 12.54)]. We checked assumptions for the PCA using the Kaiser-Meyer-Olkin measure of Sampling Adequacy and Bartlett’s test of the *REdaS* package in R (Supplemental Information) (46). The Kaiser-Meyer-Olkin measure of Sampling Adequacy (KMO =0.790) ensured that the proportion of variance among our variables, that might have been caused by an underlying factor, was large enough to be meaningful. The Bartlett’s test of sphericity [X2 (10) =608.63, *p*<0.001] indicated that the correlation matrix was not equal to an identity matrix.

We subsequently conducted the PCA using the *prcomp* function of the *stats* package (47) and the *hornpa* package (48) for Horn’s parallel analysis in R (31). The resulting screeplot of the PCA suggested a one-component solution (Supplemental Figure 4 and Supplemental Table 2). Parallel analysis, in which components are retained if the associated eigenvalue is bigger than the 95^th^ of the distribution of eigenvalues derived from a random dataset, confirmed this solution. The first component accounted for 54% of the total variance validating that all items were targeting the same underlying construct. Item loadings are additionally displayed in Supplemental Table 2 showing that all items represented an underlying construct, i.e. information seeking. Importantly, the total variance explained by the first component was higher than what could have been obtained by chance.

We obtained a Cronbach’s Alpha of *α*=0.772 for the internal consistency of our questionnaire which is considered reliable in this context (49). Finally, we measured test-retest reliability using the Pearson’s Correlation Coefficient (r(302)=0.585, *p*<0.001) suggesting a substantial test-retest reliability of our newly developed scale (50).

**Supplemental Figure 4.**
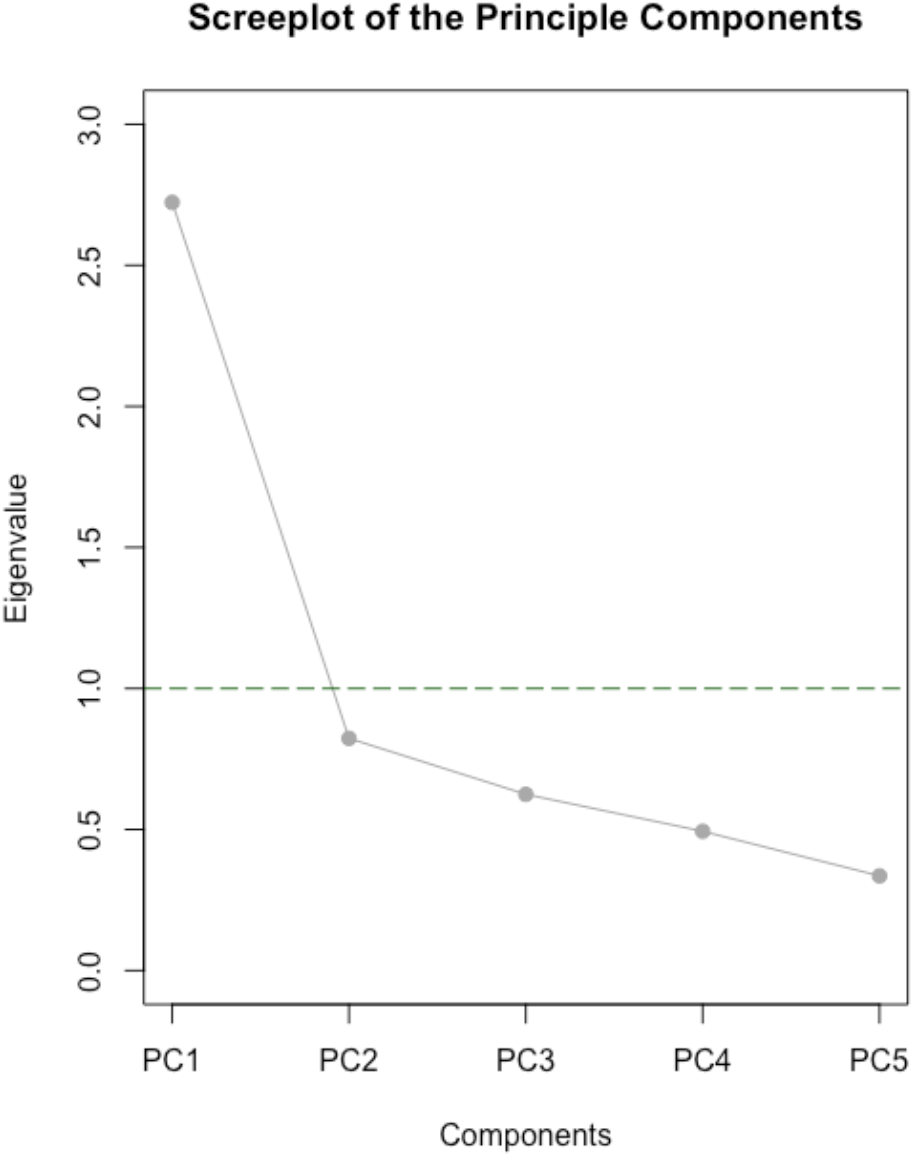
Screeplot of the Principal Component of the Covid-19 Information-seeking questionnaire. The green line indicates Eigenvalue =1. Abbreviations: PC: Principal Component.

**Supplemental Table 2.** Principal Component Analysis (PCA) loadings of the Covid-19 information-seeking questionnaire.

| Item | PC1 |
| --- | --- |
| Item 1 | 0.487 |
| Item 2 | 0.473 |
| Item 3 | 0.370 |
| Item 4 | 0.398 |
| Item 5 | 0.494 |

*Note.* We extracted one component (i.e. eigenvalue >1) using PCA, which explained 54% of the total variance.

### Decrease in information seeking from during the first pandemic wave to after the ease of lockdown

To show the robustness of our result indicating a decrease in average information seeking from T1 to T2 we conducted an additional permutation test to compare the observed difference in means 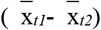 to a reference null distribution (Supplemental Figure 5 below). The observed difference in mean information seeking of T1 and T2 was significantly lower than the null distribution (*p*<0.001), confirming the decrease in information seeking from T1 to T2.

**Supplemental Figure 5.**
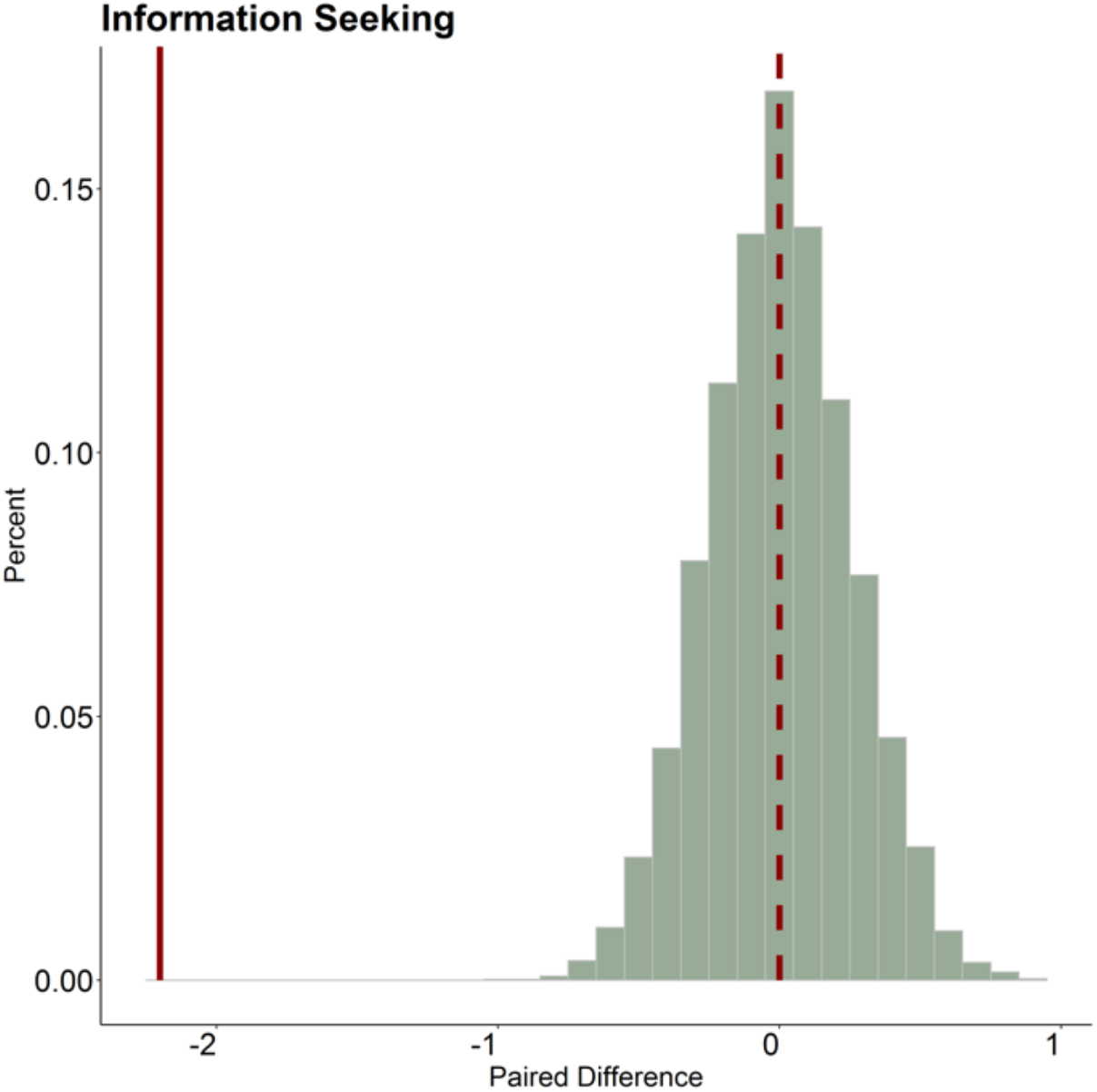
Histograms of the null distributions of the mean differences in information seeking scores between T1 and T2 (*N*=20,000 permutations). The solid red lines represent the observed difference in means while the dashed red line represents the mean of permutation distribution. The observed ratio was significantly smaller than the permutation realizations of the ratio, which provides evidence for a decrease in information seeking from T1 to T2 (*p*<0.001).

**Supplemental Table 3.**
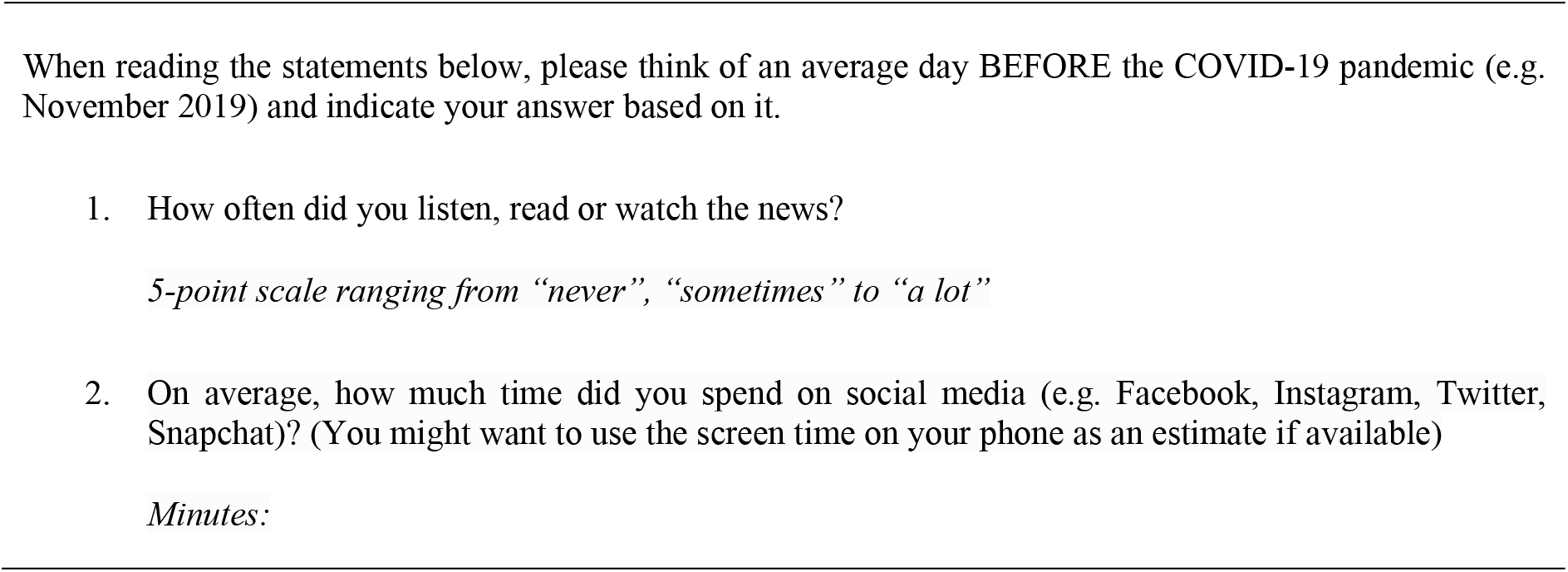
Items measuring baseline news and social media consumption.

### Adherence to pandemic-related governmental guidelines

We wanted to investigate to which degree participants adhered to guidelines advocated to prevent the further spread of Covid-19. Items were collected from the NHS and UK government website and are shown in the Supplemental Table 4 below.

**Supplemental Table 4.**
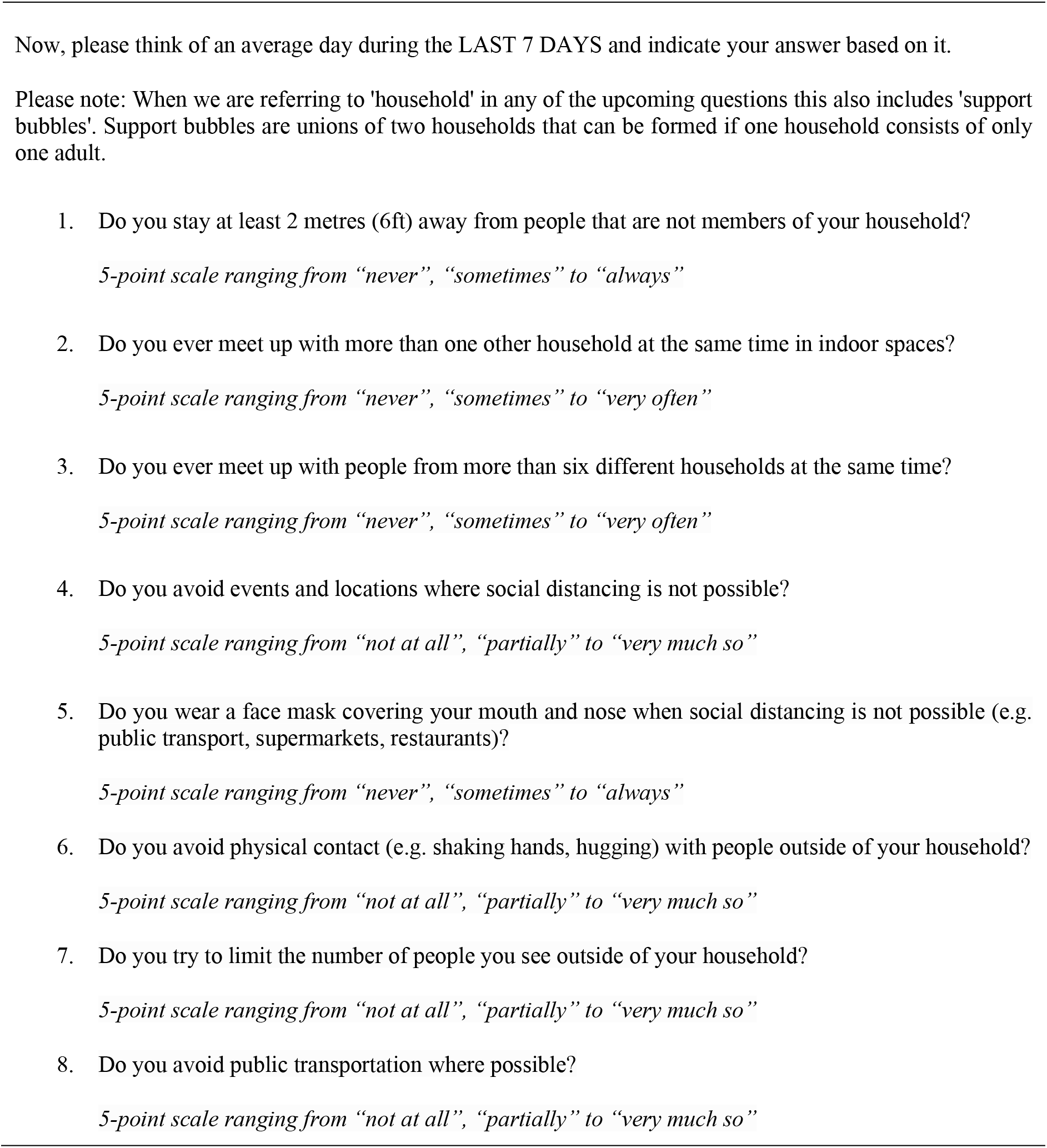
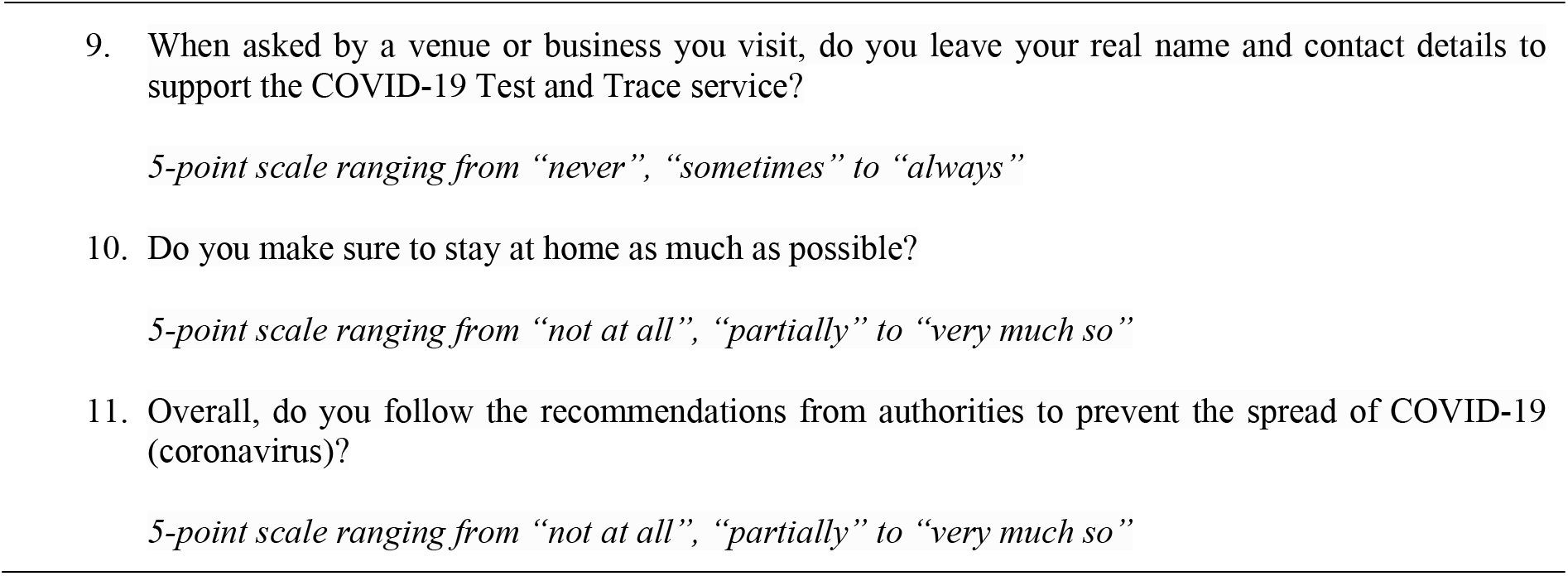
Items measuring guideline adherence.

### Mediation analyses

To test the robustness of our hypothesized mediation model (investigating the association between OC symptoms, information seeking and guideline adherence) and control for correlation effects, we ran two additional mediation models. The results of our hypothesized model were replicated when using information seeking at T2 as a mediator (path *a*: *β*=0.18, *p*<0.001; path *b*: *β*=0.27, *p*<0.001; path *ab*: *β*=0.05, *p*<0.001; path *c’*: *β*=0.09, *p*=0.138). Thus OC symptoms as well as information seeking were associated with guideline adherence at T2 but the relationship between OC symptoms and guideline adherence was mediated by information seeking. The effect of OC symptoms on guideline adherence became non-significant when controlling for information seeking.

The additional model to control for directionality by reversing OC symptoms and information seeking did not show a significant mediation effect (*β*= 0.02, *p*= 0.0625; path *ab*) whereas the significant association between information seeking and OC symptoms (*β*= 0.21, *p*<0.001; path *a*) as well as information seeking and guideline adherence (*β*= 0.15, *p*= 0.003, path *c; β*=0.13, *p*= 0.020; path *c’*) prevailed. This shows that our hypothesized model uniquely explains the relationship between OC symptoms, information seeking and guideline adherence.

